# Effects of different vestibular implant stimulation paradigms on balance and gait variability in people with severe bilateral vestibulopathy: a triple-blinded randomised cross-over study

**DOI:** 10.1101/2025.10.17.25338057

**Authors:** Meichan Zhu, Rik Marcellis, Paul Willems, Bernd Vermorken, Stan van Boxel, Joost Stultiens, Miranda Janssen, Jona Beckers, Benjamin Volpe, Gautier Grouvel, Angélica Pérez Fornos, Nils Guinand, Elke Devocht, Kenneth Meijer, Raymond van de Berg, Christopher McCrum

## Abstract

**Objectives:** The vestibular implant is a neuroprosthesis that may offer a promising treatment for patients with severe bilateral vestibulopathy (BVP). This study explored the impact of different vestibular stimulation modes on gait and balance outcomes. It was expected that stimulation modes incorporating head motion-modulated input (modes A and B) would result in more favourable gait and balance outcomes compared to non-modulated baseline stimulation or no stimulation.

**Methods:** The VertiGo! trial is a randomized controlled crossover study. The trial includes nine participants with severe BVP who received a vestibular implant. In this triple-blinded sub-study, balance and gait variability were assessed during four testing weeks, the first providing a reference (no stimulation) followed by three weeks each with four days of stimulation: (A) baseline stimulation with head motion-modulation; (B) reduced baseline stimulation with head motion-modulation, and (C) baseline stimulation without modulation. Participants walked at different walking speeds (0.6, 0.8, and 1.0m/s) on an instrumented treadmill integrated in a 6-degree of freedom motion platform. Different walking conditions were evaluated (unperturbed, three levels of mediolateral platform sway and darkness). Coefficients of variation of spatiotemporal step parameters were analysed using 3D motion capture. Participants also completed the Mini-Balance Evaluation Systems Test (Mini-BESTest) once during the reference week and twice (day 1 and day 4) of each stimulation period.

**Results:** There were no clear indications across the participants that stimulation modes A and B (compared to either mode C or no stimulation) were uniformly beneficial for either step time CoV after three days of stimulation or Mini-BESTest scores after four days of stimulation (though some individuals did demonstrate this pattern). Mini-BESTest scores significantly improved between day one and day four of stimulation.

**Conclusions:** Four days of VCI stimulation appears to affect gait variability and balance (as measured by the Mini-BESTest) but effects vary between individuals and no consistent effect of stimulation mode across participants and walking conditions was found. Balance improved from day one to day four of stimulation (without a significant learning effect over all seven clinical balance assessments), indicating the importance of habituation to vestibular implant stimulation before beneficial functional outcomes can be expected.

## Introduction

Bilateral vestibulopathy (BVP) is a condition characterized by reduced function of the vestibular nerve or labyrinth on both sides. BVP is estimated to affect between 53 and 95 million individuals across Europe and the United States^1,2^. Common symptoms include unsteadiness while walking or standing^3–6^, which worsen in low light conditions or on uneven surfaces^5–8^, and movement-induced blurred vision (oscillopsia)^3–5,9–11^. Studies of gait in BVP repeatedly report significantly increased gait variability, most often in step time, particularly at slower walking speeds^12–15^. This appears to be related to an increased risk of falls in BVP^16–18^.

Unfortunately, the prognosis for recovery of vestibular function in patients with BVP is poor^4^. While vestibular rehabilitation can lead to significant functional improvements, residual symptoms commonly persist^8,19^. Recent advancements in imperceptible noisy galvanic vestibular stimulation show potential for improving residual vestibular function, including in balance and gait, in people with partial bilateral vestibular loss^20–28^. However, for more severe or complete loss, no restorative treatment is available.

Over the last decades, multiple research groups have investigated vestibular implant prototypes to (partially) restore vestibular function in patients with BVP^29–33^. The vestibular implant is conceptually similar to a cochlear implant. It uses motion sensors to detect head movements and converts them into electrical signals. These electrical signals are then transmitted to the vestibular nerve via electrodes implanted near vestibular nerve branches that innervate the semicircular canals. Human studies have demonstrated that a vestibular implant can partially restore vestibular reflexes across low^34^, mid-^32,35^, and high-frequencies^36^. These effects contribute to improvements in dynamic visual acuity, particularly at faster head movements^37,38^ and postural responses^39,40^. Despite these promising outcomes, the extent to which these improvements translate into balance and gait remains insufficiently explored. Only one study^41^ of eight participants has investigated posture and gait in relation to a multi-canal vestibular prosthesis, using assessor-scored clinical balance and gait tests. The study did report improvement from the first measurement (before implantation) to the second measurement (6 months post-implantation) but no short- or intermediate-term analyses or quantitative gait analyses were performed.

Another consideration when evaluating the effects of a vestibular implant on balance and gait is the method of stimulation. Previous studies have primarily used electrical stimulation, delivered either as constant baseline stimulation to mimic tonic vestibular input^42–44^ or as motion-modulated stimulation that dynamically adjusts based on head movements^34,45^, aiming to replicate natural vestibular signals. There is heterogeneity in responses to vestibular prosthetic stimulation in previous studies^32,34–37,39–41,43,44,46^ and while motion-modulated stimulation seems to be more effective in general^34,35^, the effects of different modes of vestibular stimulation on balance and gait have not been studied.

To address these gaps, this study aimed to explore the effects of distinct vestibular implant stimulation paradigms in relation to each other and to a reference (no stimulation) condition on gait variability and dynamic balance in participants with severe BVP implanted with an investigational vestibular implant. It was expected that stimulation modes incorporating head motion-modulated input would result in more favourable gait and balance outcomes (i.e., reduced variability and higher Mini-BESTest scores) compared to non-modulated baseline stimulation or no stimulation.

## Methods

### Study design

The experiments and analyses presented here concerns a triple-blinded (patient, balance/gait assessor, analysing researchers) sub study of a single-centre, patient-blinded randomized controlled crossover trial (the VertiGO! Trial) performed in the Maastricht University Medical Centre+ (MUMC+). The VertiGO! Trial protocol was approved by the local medical ethical committee (NL73492.068.20/ METC 20–087), was registered at clinicaltrials.gov on 9^th^ June 2021 (NCT04918745), has an associated protocol publication^47,48^ with preliminary analysis plans and was conducted in accordance with the Declaration of Helsinki. It assessed the safety and efficacy of vestibular implant stimulation in participants with bilateral vestibulopathy and severe sensorineural hearing loss. Recruitment started on 01 July 2021 and ended on 11 June 2024. The measurements described here were conducted from 2023 to 2024. There was no patient and public involvement in the design, conduct or reporting of the trial. Note that some reports of other outcomes of the VertiGO! Trial have been published elsewhere^31,49–51^. This study is reported in accordance with the CONSORT reporting guidelines^52^. Regarding potential harms, all serious adverse events (SAEs), adverse events (AEs) or any other undesired side effects (e.g. psychological burden) were collected. Participants were asked in person about any AE at the beginning of every study visit. Since this report focuses only on the gait and balance outcomes, we do not report any SAEs or AEs not related to, or occurring during, these measurements. A full account of all SAEs and AEs will be reported in the primary trial report concerning the primary outcome and safety outcomes of the trial, which is currently in preparation.

### The vestibular implant

The details of the vestibulocochlear implant (VCI) and the surgical procedures have been described comprehensively elsewhere^47^. Briefly, the multi-canal VCI (provided by MED-EL, Innsbruck, Austria for use within an investigational device system; Figure 1) consists of three vestibular electrode leads (each with one electrode) and a cochlear electrode array containing nine electrodes positioned within the cochlea. Each vestibular electrode is placed in the ampulla of each semicircular canal. The vestibular target nerves were the lateral ampullary nerve (LAN), superior ampullary nerve (SAN), and posterior ampullary nerve (PAN). Preoperative and intraoperative imaging were utilized to optimize electrode placement and confirm positioning within 1.5 mm of the ampulla^53^. Head motion is detected using two inertial measurement units integrating gyroscopes and accelerometers^54^ housed in the coil. The audio-motion processor (AMP), worn around the patient’s neck, contains an audio processor unit and includes a base-unit, which has been designed as a research platform to provide a user interface to select various experimental setups, and to connect to the research software (AmpFit). The software packages and AMP were also provided by MED-EL (*Innsbruck*, *Austria*)^54^. The recorded motion signals are continuously transmitted to the implanted vestibular stimulator, where they are converted into pulsatile electrical stimulation patterns delivered to the vestibular nerve via the implanted electrodes.

**Figure 1.**
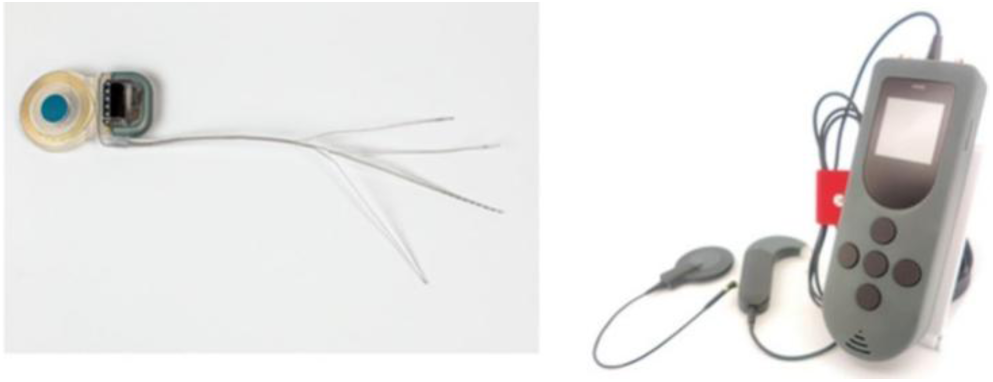
Left: A multichannel VCI was developed for the investigators using a modified cochlear implant (MED-EL, Innsbruck, Austria)^54^. It includes three vestibular electrodes designed for implantation in the three semicircular canals near the corresponding ampullary nerves, along with an array of cochlear electrodes. Right: The Audio-Motion Processor (AMP).

### Participants

Nine participants with BVP and severe sensorineural hearing loss (in the ear to be implanted), implanted with the vestibular implant were included in this study. A tenth participant was also included in the study and implanted bilaterally with vestibular implants but this participant was not assessed on the gait outcomes and is therefore not considered in these analyses. A complete justification of the sample size with regards to the primary trial outcome is provided in the protocol^47^ (section 2.5 Sample Size Calculation). Clinician study team members identified candidates and obtained written informed consent to complete further in-person vestibular testing. Participants were selected for cochlear and vestibular implantation based on the recently established consensus criteria for vestibular implantation^55,56^, meeting the following inclusion requirements: 1. Age ≥18 years; 2. Presence of disabling symptoms such as postural imbalance and/or oscillopsia; 3. Bilateral vestibulo-ocular reflex (VOR) dysfunction, demonstrated by at least one test fulfilling criterion A, with remaining tests meeting criterion B (see Table 1 in Vermorken, et al. ^47^), as defined in the vestibular implantation criteria statement^55,56^; 4. Onset of BVP after the age of two years; 5. Documented peripheral vestibular dysfunction; 6. Severe to profound sensorineural hearing loss in the ear intended for implantation, consistent with Dutch cochlear implant (CI) candidacy guidelines^57^; 7. Post-lingual onset of profound hearing loss, occurring after the age of four years. Those who were unable or unwilling to stop taking anxiety or depression medication for the week prior to the measurements were not eligible to participate. Characteristics of the included participants with a VCI (six males, three females) are shown in Table 1. As in previous publications, we refer to each participant by VCI followed by their number.

**Table 1.** Characteristics of VCI participants.

| NO. | BVP aetiology | Duration of BVP symptoms (years) | Year implanted | Implant side |
| --- | --- | --- | --- | --- |
| VCI-1 | DFNA-9 | 7 | 2021 | R |
| VCI-2 | Auto-immune (CREST) | 21 | 2021 | R |
| VCI-3 | DFNA-9 | 30 | 2022 | L |
| VCI-4 | DFNA-9 | 10 | 2022 | R |
| VCI-5 | Idiopathic | 4 | 2022 | R |
| VCI-6 | Ménière | 25 | 2022 | R |
| VCI-7 | DFNA-9 | 6 | 2022 | L |
| VCI-8 | Skull base fracture | <1 | 2023 | R |
| VCI-9 | Skull base fracture | <1 | 2023 | R |

BVP: bilateral vestibulopathy, VCI: vestibulocochlear implant, CREST: Calcinosis, Raynaud's phenomenon, Esophageal dysfunction, Sclerodactyly, and Telangiectasias.

### Schedule

As described in our protocol, following inclusion in the trial, participants were scheduled for VCI surgery. After surgery, they followed the regular cochlear implant rehabilitation program, attended one postoperative control visit, and had four hours of acute vestibular implant testing across three sessions. Once these periods were complete, a four-day vestibular implant fitting period, one day of reference testing and three periods of four days (three-period crossover design) of VCI stimulation were scheduled. The reference testing day was designed to assess the participants’ vestibular performance without vestibular stimulation. Each VCI stimulation period was used to test one of the three VCI stimulation modes. Specific details on the stimulation modes have been previously described in detail^31,49^. Briefly, for each electrode, the dynamic range in terms of current amplitude was determined. The dynamic range was defined as the range between the activation threshold (i.e. the lowest stimulation with perception and/or VOR), and the upper comfortable limit (i.e. the highest stimulation current without discomfort and/or facial nerve stimulation). Motion-modulated stimulation was applied by modulating a baseline stimulation within the dynamic range. Baseline stimulation refers to a continuous, constant-amplitude electrical pulse signal that can enable up- and down-modulation, and therefore bidirectional motion encoding. Two motion-modulated modes were assessed, one with the baseline at 50% of the dynamic range (Mode A), and one with the baseline at threshold level (Mode B) enhancing the encoding of excitation at the expense of inhibition encoding (Figure 2A and 2B). Baseline-only stimulation (Mode C) delivered a constant-amplitude electrical pulse at 50% of the dynamic range with no modulation in response to head movements (Figure 2C). After enrolment, the order of the modes was randomised between participants by one team member, using a Latin square. The order was concealed from the patients, gait researchers performing the measurements and from the researchers performing the graphing and analyses, by using a code letter (X, Y, Z) on any documentation referring to the mode. This blind was removed after the results section of the manuscript was drafted. All three electrodes were activated in all stimulation modes.

**Figure 2.**
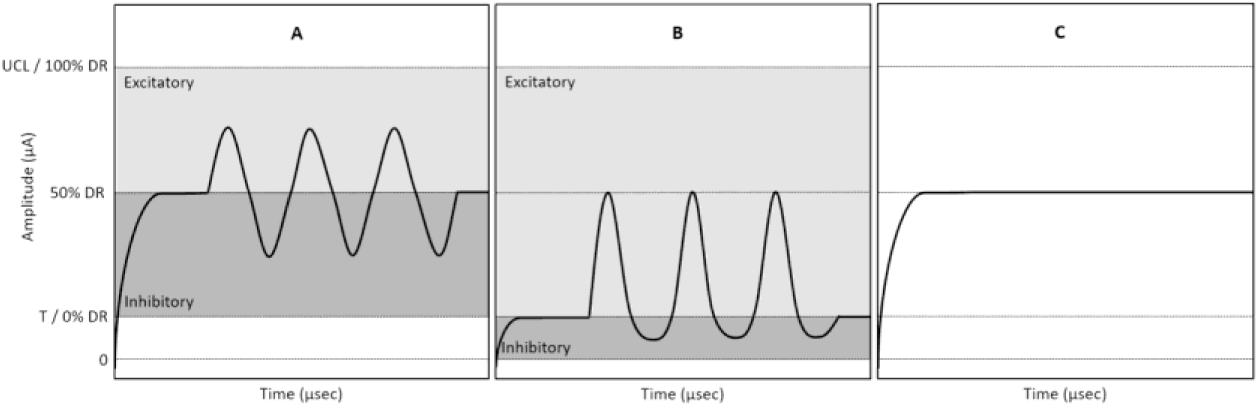
Three modes of stimulation by the VCI, adapted from Vermorken, et al. ^47,48^. (A) Motion-modulated stimulation with baseline stimulation at 50% of dynamic range (DR; range from the perceptual threshold [T] to the upper comfortable limit [UCL]). (B) Motion-modulated stimulation with reduced baseline stimulation at T. (C) Baseline stimulation (no modulation) at 50% of DR. Baseline stimulation is given as a constant-amplitude electrical pulsatile signal in all three stimulation modes. Excitatory modulation is shown in light grey and inhibitory modulation is shown in dark grey.

### Gait analysis setup and procedures

The gait measurements were conducted on the reference testing day and on the third day of each stimulation period. Measurements were conducted using the Computer Assisted Rehabilitation Environment Extended (CAREN; Motek Medical B.V., Amsterdam, The Netherlands) which consists of a dual-belt force plate-instrumented treadmill (sampling rate: 1000 Hz) mounted on a 6 degrees of freedom motion platform, a motion capture system with 12 infrared cameras (100 Hz; Vicon Motion Systems, Oxford, UK), and a 180-degree curved screen onto which a virtual environment (static city-style) is projected. The motion capture system tracked six retroreflective markers that were attached to anatomical landmarks (C7, sacrum, left and right trochanters, left and right hallux). A safety harness fastened to an overhead frame was used for all measurement sessions.

The first session consisted of familiarization trials, during which all participants began by walking at three different speeds. In our previous research, we found that participants with BVP had more difficulty walking at slower speeds (e.g., 0.4-0.8m/s) compared to faster speeds (e.g., above 1m/s)^13^. Based on these findings, three walking speeds (0.6m/s, 0.8m/s and 1.0m/s) were selected for this study. The entire procedure consisted of six sessions. The first session involved familiarization and unperturbed walking. The second session included unperturbed walking at the three selected speeds, with each speed lasting 3 minutes, for a total of 9 minutes. The third, fourth and fifth sessions each introduced pseudorandom mediolateral platform sway perturbations of increasing maximum displacements (sway1, sway2 and sway3; see below). The final session consisted of unperturbed walking in darkness, using the same speeds and durations as the standard unperturbed condition. If a participant felt uncomfortable progressing to the next speed, or if the CAREN operator or physician deemed it unsafe or impractical, the participant continued walking at the current speed. Following that, participants were given sufficient time to recover before proceeding with the measurements. Each measurement ensured a minimum of 60 recorded strides (120 steps) per speed.

The pseudorandom mediolateral platform sway perturbations were based on the perturbation presented by McAndrew, et al. ^58^ which we modified for use with people with BVP, as described in our previous work^15^, and have been shown to significantly increase gait variability^15^. These perturbations are visualised in Figure 3 with the formula included in the figure legend. In the darkness condition, the room lights, the virtual environment and strobe lights of the two motion cameras in front of the treadmill were turned off. In addition, participants were provided with sunglasses with lenses with visible light transmission category 3 (8-18%) with 100% UV protection. This resulted in a very dark environment measured between 0 to 1 Lumen (measured with the Lightmeter LM10, Fauser, Munich, Germany).

**Figure 3:**
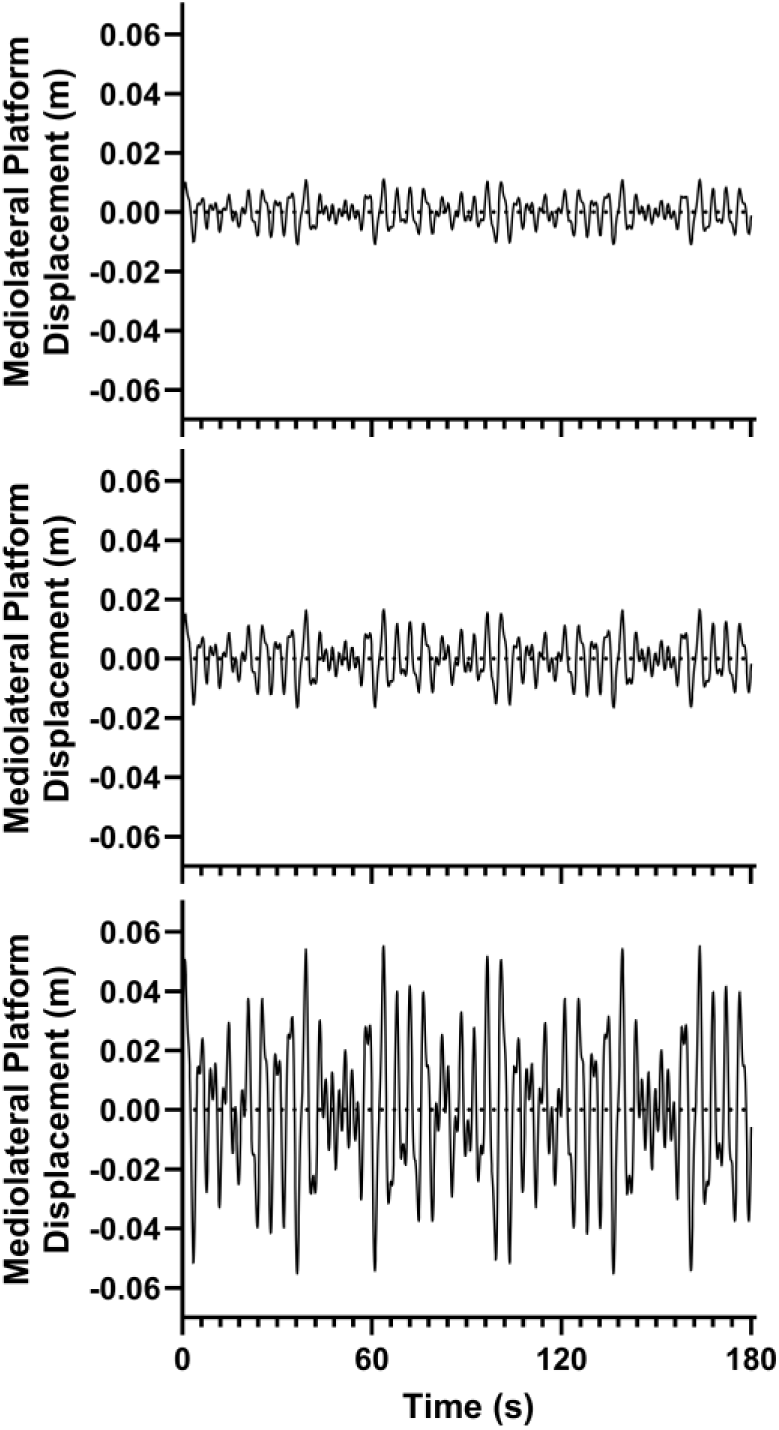
Mediolateral platform displacement generated by the final equation and scaling factors (top panel: sway 1 - scaling factor A=0.005; middle panel: sway 2 - scaling factor A=0.0075; bottom panel: sway 3 - scaling factor A=0.025 over a 180s trial. The equation used to modulate the platform position was: *D*(*t*) = *A* · [0.5 · sin(0.16 · 2*πt*) + 0.8 · sin(0.21 · 2*πt*) + 0.7 · sin(0.24 · 2*πt*) + 0.5 · sin(0.49 · 2*πt*)] where D(t) was the translation distance [m], A was a scaling factor and t was time [sec].

Regarding the gait outcomes, a low pass second order Butterworth filter (zero-phase) with a cut-off frequency of 12 Hz was used to filter marker tracks. A combined force plate (50 N threshold) and foot marker data^59^ method was used to determine foot touchdown and toe-off (as previously described in McCrum, et al. ^13^). For the final 100 steps of each trial, the means, standard deviations and coefficients of variation (CoV=SD/Mean*100%) of step time (time from touchdown of one foot to touchdown of the contralateral foot), step length (anteroposterior distance between the hallux markers at foot touchdown), double support time (time spent with both feet on the ground) and step width (mediolateral distance between the hallux markers at foot touchdown) were calculated.

### Clinical Balance Assessment

Balance assessments took place once on the reference testing day and on the first and fourth day of each stimulation period. The Mini-BESTest^60^ was used in this study to assess balance which evaluates dynamic balance across four subcomponents (anticipatory postural adjustments, reactive postural control, sensory orientation and dynamic gait) and is feasible^61^ and recommended as a comprehensive balance assessment^62^ for people with BVP. It comprises 14 items, each scored on a 3-level ordinal scale of 0 (severe = unable to perform), 1 (moderate) and 2 (normal performance) with a maximum score of 28. The Mini-BESTest was administered and rated by a trained member of the research team following the standardised procedure^60^.

### Analysis and statistics

Due to the small sample size and the number of experimental factors (three walking speeds, three walking conditions, three stimulation modes plus reference mode), it was not possible to statistically analyse these relationships’ impact on the gait outcomes. Instead, individual data points and medians were analysed and compared visually, to explore general effects of each factor and the impact of individual differences in responses. For the Mini-BESTest, however, a linear mixed model with stimulation mode, repetition number (1 or 2 within each stimulation period) and Mini-BESTest number (i.e., test number between 1 and 7; 7 tests in total: 1 during reference and 2 per stimulation mode) as fixed effects and participant number and stimulation mode as random intercepts converged and was used to analyse the total score of Mini-BESTest data (main effects only without interactions) using SPSS (v.27, IBM SPSS Statistics for Windows, NY). A p-value <0.05 was considered statistically significant. GraphPad Prism 9 (GraphPad Software, San Diego, USA) was used for all visualisations.

### Protocol Deviations

In the original protocol, it was only planned to analyse unperturbed gait and perturbed gait using the sway1 and sway2 perturbations. During measurements with the first two participants, it became evident that the protocol’s physical demands fell below the anticipated thresholds for most participants. This reduced challenge resulted in less need for inter-trial recovery periods, generally more capacity on the part of the participants to complete more trials than expected and more time within the allotted time slot of the measurement week. Therefore, we added the sway3 and darkness conditions in order to increase the challenge and specificity of the tasks to the commonly reported symptom-inducing environmental situations of people with BVP (uneven ground and dim light)^7,18,63^. These additional tasks were only conducted when the planned tasks had been completed, when the participants felt able to walk more, and when there was still time remaining. As a result, fewer participants completed these trials and therefore these trials should only be interpreted in an exploratory manner.

## Results

### Participants and Completed Walking Conditions

Seven participants completed all pre-planned walking conditions (unperturbed gait and gait with the perturbations sway1 and sway2) in all reference and stimulation periods. Two participants additionally completed the sway3 and darkness walking conditions in all reference and stimulation periods. The walking condition completion status of all participants is presented in eTable 1 in the supplement. Regarding the MiniBEST, only one participant missed one of the two tests during the stimulation week due to illness, which was recorded as an AE due to the nausea potentially being related to the stimulation. No other AEs or SAEs occurred during the time of the measurements covered here. AEs and SAEs that occurred around the period of the surgery or fitting or during tests related to other outcome measures will be reported in the upcoming primary report of the trial’s primary and safety-related outcomes.

### Step time and step time variability

#### Unperturbed gait and gait during sway1 and sway2

Step time means and CoV consistently reduced with increasing walking speeds across all walking conditions (Figure 4; e.g., for step time CoV, medians of 4.8, 4.2 and 3.2 [unperturbed], 5.3, 4.2 and 3.5 [sway1] and 5.3, 3.3 and 3.2 [sway2] for 0.6, 0.8 and 1.0m/s, respectively). Step time means and CoV did not show clear, consistent changes as a result of the sway1 and sway2 perturbations (square and triangle plots in Figure 4). Step time means and CoV did not show clear, consistent changes as a result of the stimulation modes, compared to the reference mode, though responses appeared to vary across individuals (Figure 4). For example, VCI-7 and VCI-8 do seem to show reduced step time CoV in all stimulation modes compared to the reference mode, but other participants seem to increase in step time CoV during one or more stimulation modes (e.g., VCI-1, VCI-2, VCI-3) whereas with VCI-4 and VCI-5, it appears to vary more based on the walking condition than the stimulation mode.

**Figure 4.**
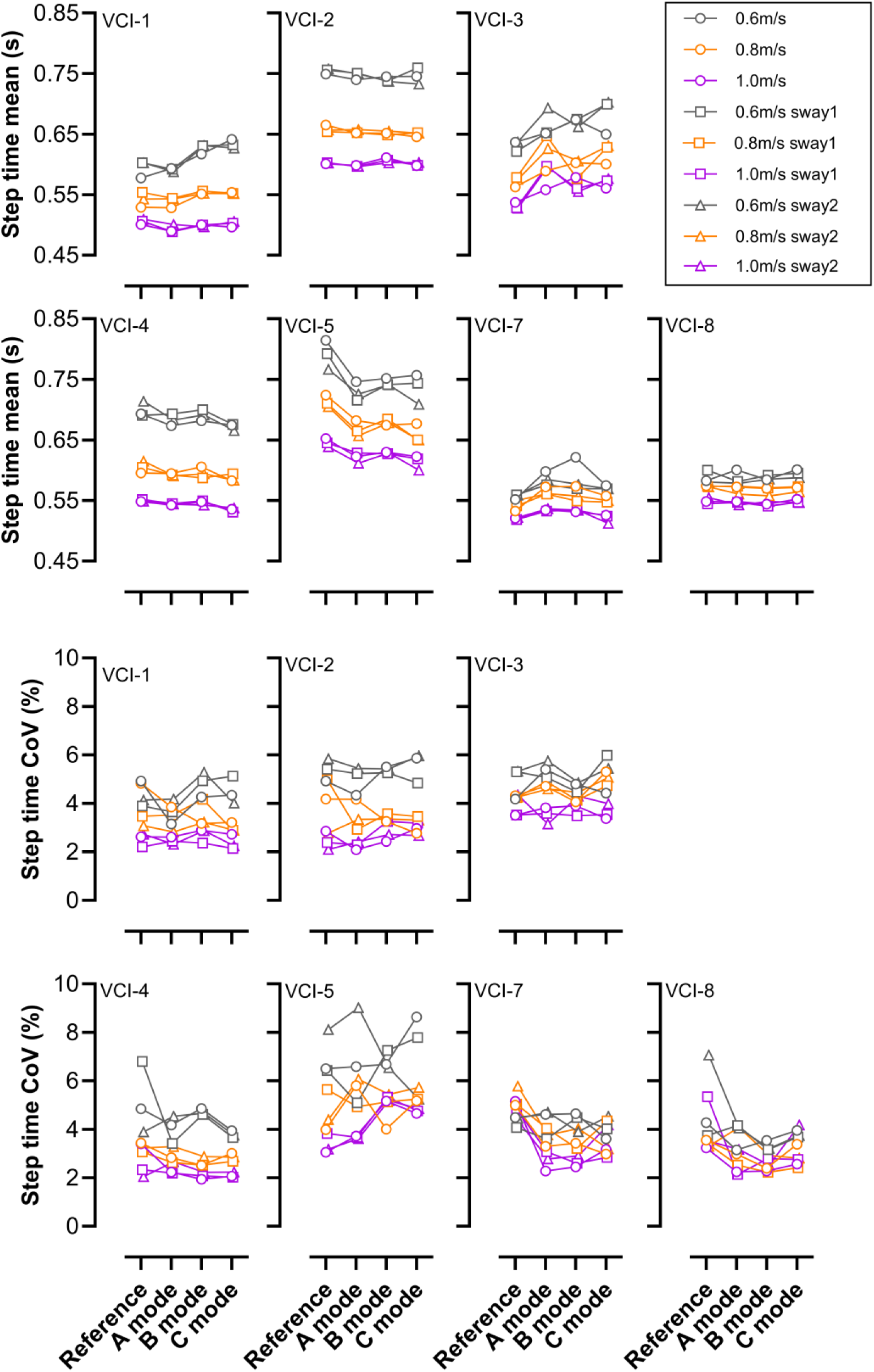
Step time (Mean and CoV) of 7 VCI participants obtained during different walking conditions (unperturbed, sway1 and sway2; sway1: pseudorandom mediolateral platform sway with scaling factor A=0.005; sway 2: pseudorandom mediolateral platform sway, scaling factor A=0.0075 [see Figure3]) under different stimulation modes (Reference, A, B and C; see Figure 2). Each data point is derived from the final 100 steps of each condition.

### Unperturbed gait and gait during sway3 and darkness

Step time means and CoV typically reduced with increasing walking speed in VCI-5 and VCI-7 across unperturbed and sway3 conditions (Figure 5; e.g., for step time CoV, medians of 5.5, 4.5 and 4.1 [unperturbed] and 8.2, 7.5 and 6.0 [sway3] for 0.6, 0.8 and 1.0m/s, respectively). Sway3 consistently led to increased step time CoV across most stimulation modes (Figure 5). While there is quite some variation in stimulation mode-related changes in the step time CoV, stimulation mode A appears to have consistently led to lower step time CoV during sway3 compared to the reference mode for these two participants (Figure 5; medians of 6.3, 5.3 and 4.3 [mode A] and 8.2, 7.5 and 6.0 [reference] for 0.6, 0.8 and 1.0m/s, respectively).

**Figure 5.**
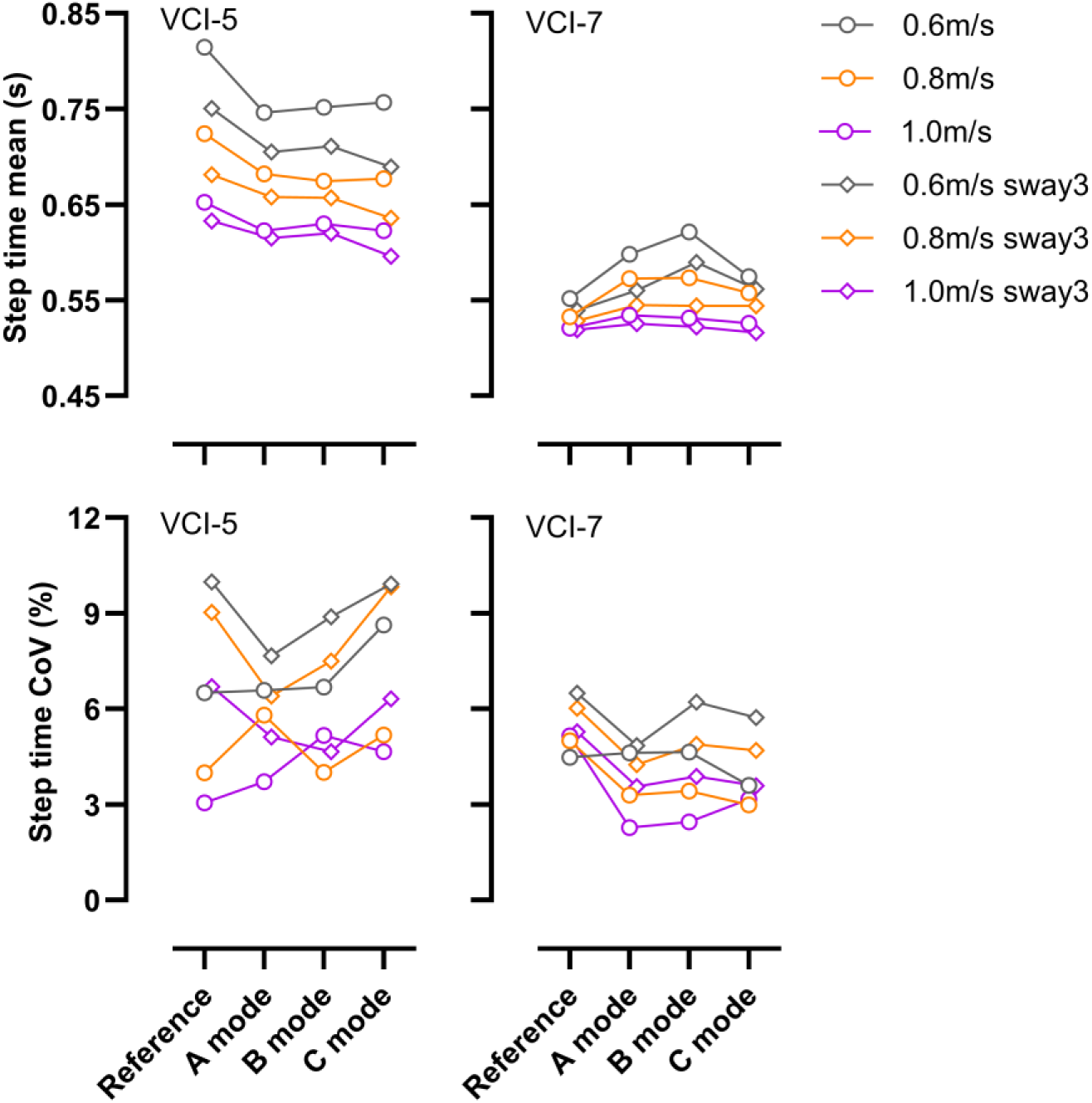
Step time (Mean and CoV) of 2 VCI participants obtained during different walking conditions (unperturbed, sway 3: pseudorandom mediolateral platform sway, scaling factor A=0.025 [see Figure3]) under different stimulation modes (Reference, A, B and C; see Figure 2). Each data point is derived from the final 100 steps of each condition.

Step time means typically reduced with increasing walking speed in VCI-5 and VCI-7 across unperturbed and darkness conditions, but step time CoV during darkness did not show as consistent an effect of walking speed (Figure 6; e.g., for step time CoV, medians of 5.5, 4.5 and 4.1 [unperturbed], 7.7, 6.5 and 5.7 [darkness] for 0.6, 0.8 and 1.0m/s, respectively). Darkness consistently led to reduced step time means and increased step time CoV across stimulation modes in VCI-7 but in VCI-5 only during 0.8 and 1.0 m/s (Figure 6). There was no clear and consistent effect of any stimulation mode on step time CoV during darkness in these two participants (Figure 6).

**Figure 6.**
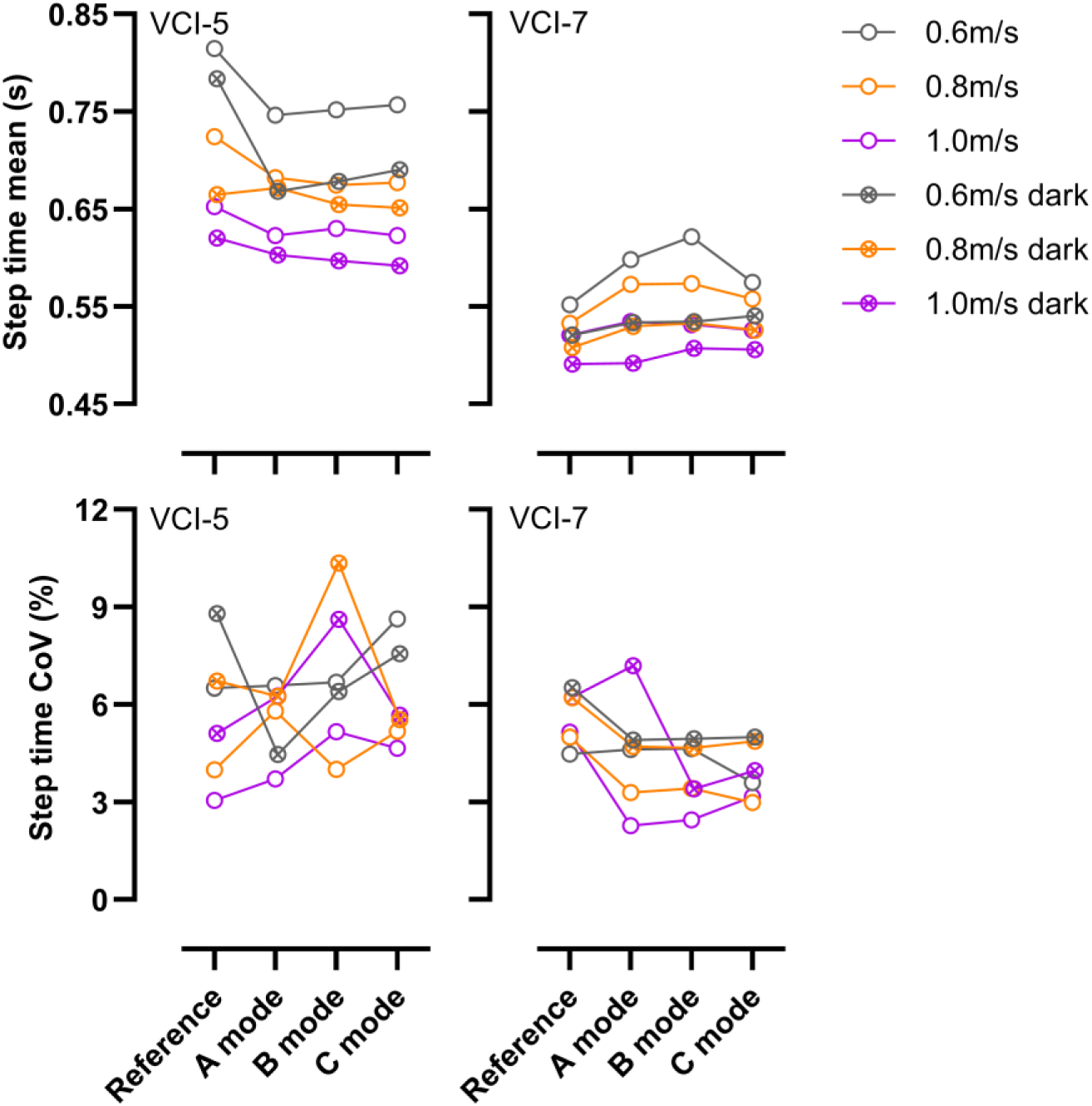
Step time (Mean and CoV) of 2 VCI participants obtained during different walking conditions (unperturbed, darkness) under different stimulation modes (Reference, A, B and C; see Figure 2). Each data point is derived from the final 100 steps of each condition.

### Step length, width and double support time

The results of the means and CoV of step length, step width and double support time are reported in the supplement (eFigures 1-9 in the supplement). Overall, similar patterns of results were seen, including a consistent effect of walking speed (except for step length CoV and double support time CoV and step width mean in darkness, as well as double support time CoV in sway3), no large effect of sway1 and sway2, larger effects of sway3 and darkness (though slightly less consistently) and no clear, consistent beneficial (i.e., lower variability) effect of any single stimulation mode across participants.

### Clinical Balance Assessment with the Mini-BESTest

In the Mini-BESTest, there was not a consistent effect of any specific stimulation mode. While at least one stimulation mode total score was higher than the reference total score for most participants, these were distributed across the different stimulation modes (Figure 7). A two-level random intercept model, accounting for random intercepts per subject and per VCI mode, revealed a repetition effect (higher score) in the Mini-BESTest when comparing the first and second assessments within each stimulation mode (F_1,26_ = 6.604, p = 0.016) with a mean difference of 1.659 (95% CI 0.331 to 2.987). However, no significant effect of VCI stimulation mode (F_3,25_ = 1.300, p = 0.297) or Mini-BESTest number (1 to 7; F_4,4_ = 2.013, p = 0.257) were observed.

**Figure 7.**
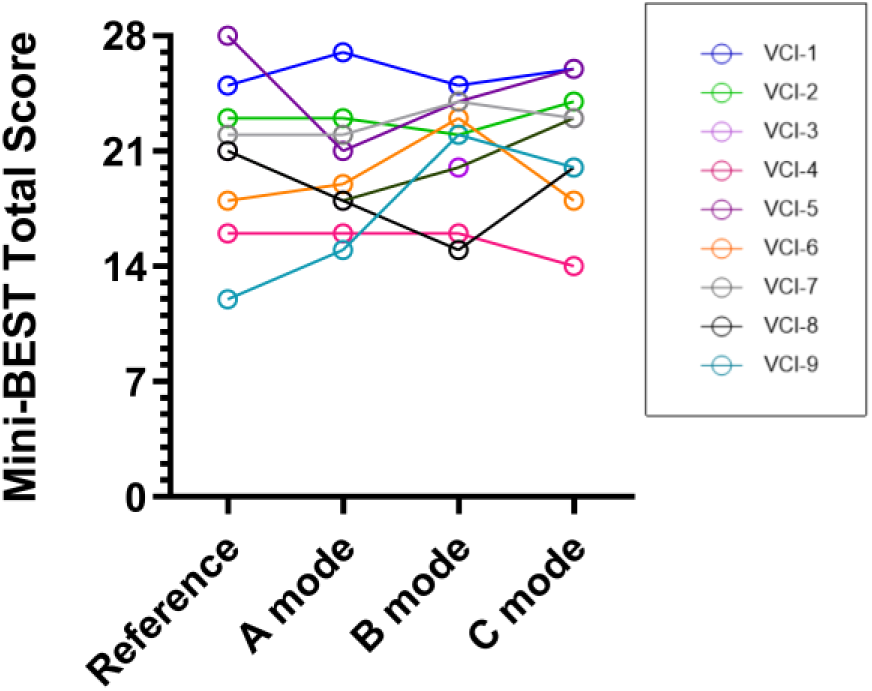
Comparison VCI participants’ (indicated by colour) Mini-BESTest results under different stimulation modes (Reference, A, B and C). For this figure, the best of the two scores in the stimulation weeks were used.

### Exploratory Analyses: Participants with and without DFNA9

The DFNA9 mutation, an autosomal dominant hereditary condition caused by mutations in the COCH gene, is known to be associated with vestibular dysfunction^64,65^. The presence of this mutation was not part of our original research questions in this study but 4 out of the 9 participants were known to carry this mutation and these participants have shown reduced responses to VCI stimulation in A, B and C stimulation modes (lower eye velocities [see Figure 3, e.g., VCI-1 and VCI-4, in van Boxel, et al. ^31^] and movement perception [unpublished observations]). Given its potential impact, we explored the potential influence of the DFNA9 mutation on the primary outcome step time CoV, as well as the Mini-BEST scores. These analyses did not reveal a consistent pattern of differences between DFNA9 carriers and non-carriers and are reported in full in eResults in the supplement.

## Discussion

This study aimed to explore the effects of distinct VCI stimulation modes in relation to each other and to a reference (no stimulation) mode on gait variability and dynamic balance in participants with severe BVP and a VCI. It was expected that stimulation modes incorporating head motion-modulated input would result in more favourable gait and balance outcomes compared to non-modulated baseline stimulation or no stimulation. However, the findings could not support this expectation, since there were no clear indications across the participants that stimulation modes A and B were uniformly beneficial for either step time CoV after three days of stimulation or Mini-BESTest scores after four days of stimulation (though some individuals did demonstrate this pattern). However, one general hypothesis of the entire VertiGO! Trial (hypothesis 3 in the protocol^47^), that outcomes measured multiple times during a stimulation period would show improvement throughout the stimulation period, was supported by the current Mini-BEST test results, since a significant repetition effect within the stimulation weeks was found, with no significant learning effect across all 7 tests.

The significant repetition effect for the Mini-BESTest results from the first test to the second test within each stimulation mode might be due to participants becoming habituated to the specific stimulation mode. This might involve habituation to misalignment that might be present between the actual head rotation and the electrically evoked vestibulo-ocular reflex since the first test occurred on the first day of stimulation and the second test occurred on the fourth and final day of stimulation. This aligns with hypothesis 3 presented in the protocol paper that outcomes were expected to improve throughout each stimulation period^47^. This change from Mini-BESTest one to two within each stimulation week, averaged across all stimulation modes, is visualised in Figure 8 and highlights this effect.

**Figure 8:**
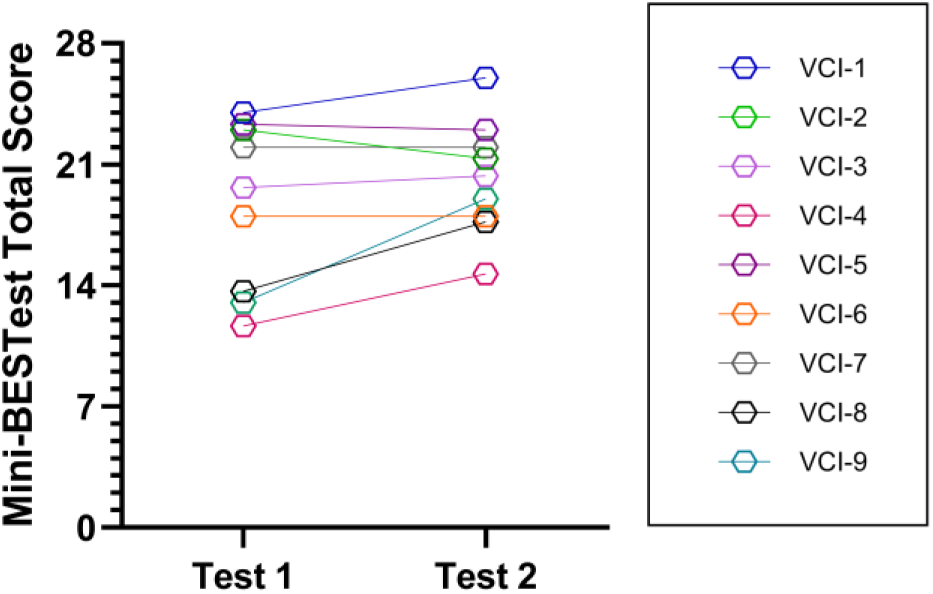
Mean of Mini-BESTest total scores from each of participants (A,B and C mode) from the first and second tests within each stimulation period (symbols) for each participant (colours).

In contrary to our hypotheses, the VCI stimulation modes did not appear to induce consistent changes in step time mean or CoV across participants. Similarly, in the Mini-BESTest, there was not a consistent effect of any specific stimulation mode and no significant learning effect over all 7 tests. With the gait measurements occurring on the third day of the stimulation period, this might have been too early to expect to observe consistent positive effects. This might be partly supported by the significant improvement in the Mini-BESTest within each stimulation period from day 1 to day 4. Experience from cochlear implant users might also support this, since they often require time to adapt to electrical stimulation before achieving optimal auditory performance^66,67^.

The inter-individual differences observed here suggest that the response to vestibular stimulation is heterogeneous and as well as the rate and extent of habituation to VCI stimulation indicated above, may depend on multiple additional factors, such as the degree of residual vestibular function, compensatory strategies (pre- and post-implantation) or individual sensitivity to sustained artificial vestibular input over days (or sensitivity to electrical stimulation in general). The absence of a uniform effect of stimulation mode on gait variability, as with other outcomes such as peak eye velocities during electrically evoked vestibulo-occular reflexes^51^, underscores the need for further investigation into personalized VCI parameter optimization, despite some outcomes showing more consistent improvements, such as peak eye velocity and VOR gain (unpublished observations). While some individuals may benefit from specific stimulation parameters, others may not experience measurable functional improvements without more prolonged habituation or tailored parameter tuning. Strong stimulation inputs may provoke overstimulation and result in similar effects to inflammation (neuritis) in the vestibular nerve, triggering compensatory neural adaptations^68^. While compensation can enhance long-term stability, the acute effects may not be beneficial or might be temporarily destabilizing, as the nervous system adjusts to the new input. Taken together, these findings and factors underscore the importance of further research on the optimisation and personalisation of stimulation parameters and physical therapy to individual thresholds, responses and compensation. A final factor that might be considered is the balance and gait ability before implantation. When comparing the step time CoV values of the current study against those of our previous study in people with BVP and healthy age-sex-matched controls who followed the same experimental gait protocol^15^ (Figure 9), the participants with VCIs in the current study, while within the range of a broader BVP sample, do tend to have slightly lower gait variability in most walking conditions, though not to the extent of the healthy participants and not to the extent that a ceiling effect of stimulation could be expected. While not directly related to gait variability, it is also worth mentioning that the participants with VCIs in the current study had slightly lower body mass index values than the participants with BVP in the previous study (participants with VCIs (n=9): mean(SD) = 25.8(3.8); previous participants with BVP (n=42): 27.7(5.3)).

**Figure 9.**
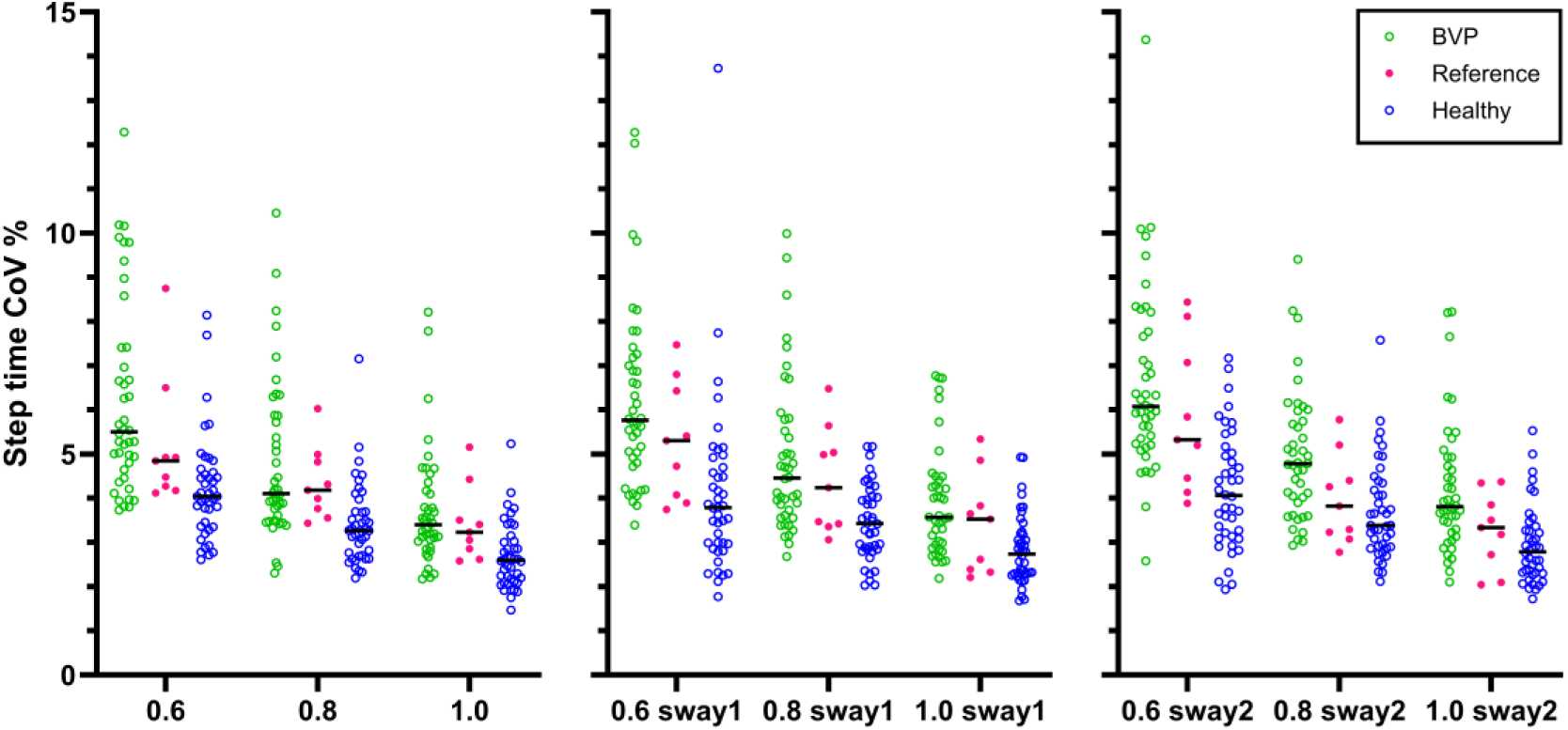
Step time CoV (individual data and group medians) obtained during different walking conditions (no sway, sway1 and sway2) at three walking speeds (0.6, 0.8 and 1.0m/s) from the VCI participants of the current study in the reference week (red, centre) compared to people with BVP (green, left) and healthy age-sex-matched control participants (blue, right) collected in our previous study^15^. Each data point is derived from the final 100 steps of each condition.

The present findings demonstrate that both step time mean and its variability, expressed as the CoV, decreased systematically with increasing walking speed across all walking conditions. This trend is consistent with previous studies^12,13,15,69,70^ lending credibility to the validity of our data. The sway perturbations (sway1 and sway2) did not lead to clearly visible effects on step time CoV. Our previous study in 42 participants with BVP and 42 age-sex-matched healthy controls did find a statistically significant effect of these same perturbations on step time CoV, but the differences were relatively small and the statistical result may have been related to the relatively large sample^15^. The small sample of the current study probably prevented us observing these small differences. However, sway3 did appear to consistently increase step time CoV in VCI-5 and VCI-7, as well as in all other participants with partial data from this condition (data not shown; see shared individual data at https://doi.org/10.17605/OSF.IO/JDUY7), suggesting that this condition imposed a greater challenge, as intended. Interestingly, stimulation mode A appeared to reduce this destabilizing effect in both participants by consistently reducing step time CoV (by 2-3%) compared to the reference mode. In the darkness condition, while step time CoV showed less consistency, it was typically higher compared to the lit condition, indicating the increased challenge of the condition. Unlike in sway3 however, there was not a clear effect on any one stimulation mode across participants VCI-5 and VCI-7. These findings together might suggest that the specific context of the walking task (i.e., managing larger physical disturbances versus coping with less visual input) may be a factor in whether VCI stimulation (and type of stimulation) is likely to improve performance. This emphasizes the complexity and task-specific nature of multisensory integration in gait, adding another factor to the considerations needing further exploration mentioned above.

Despite initial observations in other outcomes that participants carrying DFNA9 might be less responsive to the VCI (not statistically analysed), the current results do not indicate that they responded worse to VCI stimulation than non-carriers (eResults in supplement). This lack of a clear pattern may be attributed to the small sample size, the high between-individual variability or may be due to DFNA9 not particularly negatively impacting gait and balance performance. To explore this further, we visually compared the step time CoV results from the same unperturbed, sway1 and sway2 walking conditions from our previous study on people with BVP^15^, separating the group into diagnosed DFNA9 carriers and non-carriers and found that the values typically overlap (eResults in supplement). This aligns with our suggestion that the effects of DFNA9 might not particularly negatively impact gait and balance performance.

The current stage of vestibular implant research, this study, faces the inherent limitation of small sample sizes (small trials with prototype implants, invasive procedures with strict inclusion criteria^41,47^). As a result, the current conclusions are primarily based on visual observations of the data, its medians and distributions and less on comprehensive and well-powered statistical analyses. At this stage, the aim is not to generalise the findings to a broader population but rather explore the effects on this specific group of participants and guide further research and development. In this study, the interactions between all theoretically relevant parameters could not be fully explored without exceeding the acceptable burden on participants, so some gaps remain for future research to address. The current study focused on step time CoV as its primary outcome, since this is the most established gait parameter affected by BVP in the literature^12–15^. However, step time CoV does not necessarily capture all aspects of walking stability and variability and there is evidence of other parameters also showing deficits in BVP^14,71,72^. Additionally, head, trunk and upper limb kinematics were not considered in the current study due to the reduced marker set used and head stabilisation would be a parameter of interest. It is also worth noting that treadmill walking at a fixed speed may not completely represent walking in daily life in which people have the ability (and necessity) to make adjustments to their walking speed, so future investigation of overground walking tasks will be of value. Finally, since the measurements were conducted on different days, variation in performance due to factors other than the task or stimulation mode may have occurred. Once some of the factors relating to the VCI and individual differences described above have been addressed, future studies could explore the influence of the VCI on a broader range of gait variability and stability outcomes in a wider range of settings and tasks. Finally, in this trial, participants habituation to the stimulation was evaluated by confirming no nystagmus and no discomfort. However, these criteria do not necessarily mean that sensory integration of the new vestibular input is habituated or complete, as the improvement in balance from day 1 to day 4 might indicate. A more functional definition of habituation to artificial vestibular stimulation could be useful for future studies.

In conclusion, VCI stimulation over a four day period affects gait and balance but effects vary between individuals and the three stimulation paradigms did not show a clearly consistent effect across participants and walking conditions. Balance improved from day one to day four of stimulation (without a significant learning effect over all seven clinical balance assessments), indicating the importance of habituation to vestibular implant stimulation before beneficial functional outcomes can be expected. Results in two individuals while walking with large mediolateral perturbations and in darkness give preliminary indications that benefits might also be task-specific. These findings underscore the complexity of evaluating vestibular implants in the short term and emphasize the need for further research into optimisation and personalisation of stimulation paradigms and physical therapy, as well as longer-term studies.

## Supporting information

CONSORT 2025 Checklist

## Author contributions

**A.P.F.:** Conceptualization, Funding acquisition, Project administration, and Writing - review & editing.

**B. Vermorken:** Investigation, Project administration, and Writing - review & editing.

**B. Volpe:** Investigation and Writing - review & editing.

**C.M.:** Conceptualization, Data curation, Formal analysis, Funding acquisition, Investigation, Methodology, Project administration, Software, Supervision, Visualization, Writing - original draft, and Writing - review & editing.

**E.D.:** Conceptualisation, Methodology, Project administration and Writing - review & editing.

**J.B.:** Data curation, Investigation, Project administration, Resources, and Writing - review & editing.

**J.S.:** Investigation and Writing - review & editing.

**K.M.:** Conceptualization, Funding acquisition, Methodology, Project administration, Resources, Supervision, and Writing - review & editing.

**M.J.:** Formal analysis, Methodology, and Writing - review & editing.

**M.Z.:** Data curation, Formal analysis, Investigation, Project administration, Visualization, Writing - original draft, and Writing - review & editing.

**N.G.:** Conceptualization, Funding acquisition, Project administration, and Writing - review & editing.

**P.W.:** Methodology, Resources, Software, and Writing - review & editing.

**R.M.:** Conceptualization, Data curation, Investigation, Project administration, Resources, and Writing - review & editing.

**R.v.d.B.:** Conceptualization, Funding acquisition, Methodology, Project administration, Resources, Supervision, and Writing - review & editing.

**S.v.B.:** Investigation and Writing - review & editing.

## Data availability

Individual data and SPSS syntax are available at the public OSF project page: https://doi.org/10.17605/OSF.IO/JDUY7

