## Supplement for "Effects of different vestibular implant stimulation paradigms on balance and gait variability in people with severe bilateral vestibulopathy: a triple-blinded randomised cross-over study"

[1]Department of Otorhinolaryngology and Head and Neck Surgery, Division of Balance Disorders, Maastricht University Medical Centre (MUMC+), Mental Health and Neuroscience Research Institute, Maastricht, The Netherlands
[2]Department of Nutrition and Movement Sciences, Institute of Nutrition and Translational Research in Metabolism (NUTRIM), Maastricht University, Maastricht, The Netherlands
[3]Department of Physical Therapy, MUMC+, Maastricht, The Netherlands
[4]Department of Methodology and Statistics, Care and Public Health Research Institute (CAPHRI), Maastricht University, Maastricht, The Netherlands
[5]Service of Otorhinolaryngology and Head and Neck Surgery, Department of Clinical Neurosciences, Geneva University Hospitals, Geneva, Switzerland
[6]Kinesiology Laboratory, Geneva University Hospitals and University of Geneva, Geneva, Switzerland
[7]Research Center of Skeletal Muscle and Movement, Geneva University Hospitals and University of Geneva, Geneva, Switzerland
#: Joint last authors

*Correspondence:
Meichan Zhu:
Christopher McCrum:

**eResults**

**eTable 1.** The walking condition completion status of all participants (n=9)

| ID | Stimulation Period | Stimulation Mode | Unperturbed | | | sway1 | | | sway2 | | | sway3 | | | darkness | | |
|---|---|---|---|---|---|---|---|---|---|---|---|---|---|---|---|---|---|
| | | | 0.6 m/s | 0.8 m/s | 1.0 m/s | 0.6 m/s | 0.8 m/s | 1.0 m/s | 0.6 m/s | 0.8 m/s | 1.0 m/s | 0.6 m/s | 0.8 m/s | 1.0 m/s | 0.6 m/s | 0.8 m/s | 1.0 m/s |
| **VCI01** | 1 | Reference | √ | √ | √ | √ | √ | √ | √ | √ | √ | NA | NA | NA | NA | NA | NA |
| | 2 | A | √ | √ | √ | √ | √ | √ | √ | √ | √ | NA | NA | NA | NA | NA | NA |
| | 3 | B | √ | √ | √ | √ | √ | √ | √ | √ | √ | NA | NA | NA | NA | NA | NA |
| | 4 | C | √ | √ | √ | √ | √ | √ | √ | √ | √ | NA | NA | NA | NA | NA | NA |
| **VCI02** | 1 | Reference | √ | √ | √ | √ | √ | √ | √ | √ | √ | NA | NA | NA | NA | NA | NA |
| | 2 | B | √ | √ | √ | √ | √ | √ | √ | √ | √ | NA | NA | NA | NA | NA | NA |
| | 3 | A | √ | √ | √ | √ | √ | √ | √ | √ | √ | NA | NA | NA | NA | NA | NA |
| | 4 | C | √ | √ | √ | √ | √ | √ | √ | √ | √ | NA | NA | NA | NA | NA | NA |
| **VCI03** | 1 | Reference | √ | √ | √ | √ | √ | √ | √ | √ | √ | NA | NA | NA | NA | NA | NA |
| | 2 | C | √ | √ | √ | √ | √ | √ | √ | √ | √ | NA | NA | NA | NA | NA | NA |
| | 3 | B | √ | √ | √ | √ | √ | √ | √ | √ | √ | √ | √ | √ | √ | √ | √ |
| | 4 | A | √ | √ | √ | √ | √ | √ | √ | √ | √ | √ | √ | √ | √ | √ | √ |
| **VCI04** | 1 | Reference | √ | √ | √ | √ | √ | √ | √ | √ | √ | NA | NA | NA | NA | NA | NA |
| | 2 | B | √ | √ | √ | √ | √ | √ | √ | √ | √ | √ | √ | √ | √ | √ | √ |
| | 3 | A | √ | √ | √ | √ | √ | √ | √ | √ | √ | √ | √ | √ | √ | √ | √ |
| | 4 | C | √ | √ | √ | √ | √ | √ | √ | √ | √ | √ | √ | √ | √ | √ | √ |
| **VCI05** | 1 | Reference | √ | √ | √ | √ | √ | √ | √ | √ | √ | √ | √ | √ | √ | √ | √ |
| | 2 | B | √ | √ | √ | √ | √ | √ | √ | √ | √ | √ | √ | √ | √ | √ | √ |
| | 3 | C | √ | √ | √ | √ | √ | √ | √ | √ | √ | √ | √ | √ | √ | √ | √ |
| | 4 | A | √ | √ | √ | √ | √ | √ | √ | √ | √ | √ | √ | √ | √ | √ | √ |
| **VCI06** | 1 | Reference | √ | √ | √ | √ | √ | √ | √ | √ | √ | x | x | x | x | x | x |
| | 2 | C | √ | √ | x | √ | √ | x | √ | √ | x | x | x | x | x | x | x |
| | 3 | B | √ | √ | √ | √ | √ | √ | √ | √ | √ | √ | √ | √ | √ | √ | √ |
| | 4 | A | √ | √ | √ | √ | √ | √ | √ | √ | √ | √ | √ | √ | √ | √ | √ |
| **VCI07** | 1 | Reference | √ | √ | √ | √ | √ | √ | √ | √ | √ | √ | √ | √ | √ | √ | √ |
| | 2 | B | √ | √ | √ | √ | √ | √ | √ | √ | √ | √ | √ | √ | √ | √ | √ |

| | | | | | | | | | | | | | | | | | |
|---|---|---|---|---|---|---|---|---|---|---|---|---|---|---|---|---|---|
| | 3 | C | √ | √ | √ | √ | √ | √ | √ | √ | √ | √ | √ | √ | √ | √ | √ |
| | 4 | A | √ | √ | √ | √ | √ | √ | √ | √ | √ | √ | √ | √ | √ | √ | √ |
| **VCI08** | 1 | Reference | √ | √ | √ | √ | √ | √ | √ | √ | √ | x | x | x | √ | √ | √ |
| | 2 | C | √ | √ | √ | √ | √ | √ | √ | √ | √ | x | x | x | √ | √ | √ |
| | 3 | A | √ | √ | √ | √ | √ | √ | √ | √ | √ | x | x | x | √ | √ | √ |
| | 4 | B | √ | √ | √ | √ | √ | √ | √ | √ | √ | √ | √ | √ | √ | √ | √ |
| **VCI09** | 1 | Reference | √ | √ | √ | √ | √ | √ | √ | √ | √ | √ | √ | x | x | x | x |
| | 2 | C | √ | √ | √ | √ | x | x | x | x | x | x | x | x | x | x | x |
| | 3 | A | √ | √ | √ | √ | √ | √ | √ | x | x | x | x | x | x | x | x |
| | 4 | B | √ | √ | √ | x | x | x | x | x | x | x | x | x | x | x | x |

√ = walking condition completed, x = not able to start or complete walking condition, NA = not applicable since the walking condition was not planned (see protocol deviations section of main manuscript), VCI: vestibulocochlear implant; reference mode: no stimulation; A,B,C stimulation mode: (A) baseline stimulation with head motion-modulation, (B) stimulation with reduced baseline stimulation with head motion-modulation, and (C) baseline stimulation without modulation.

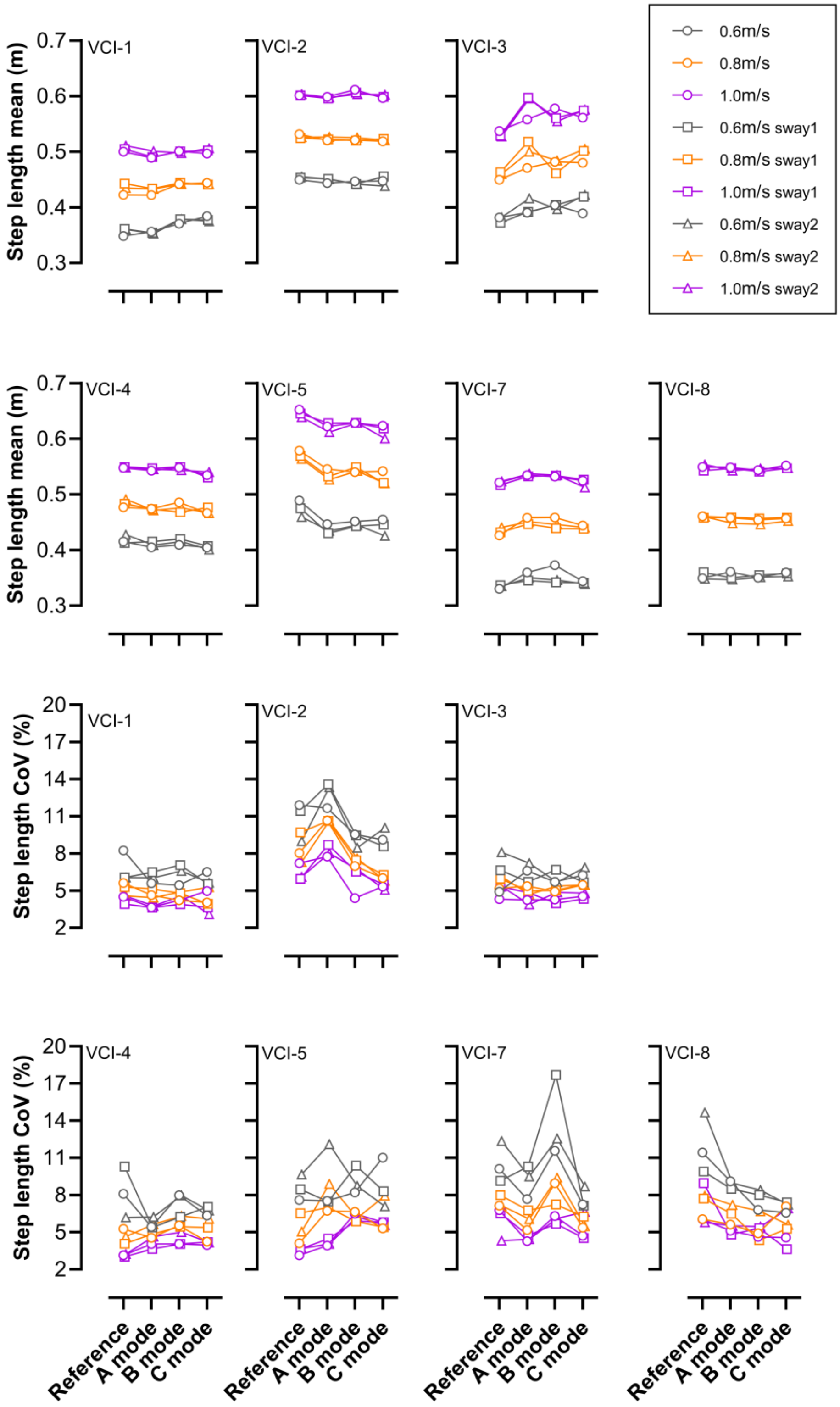


**eFigure 1**. Step length (mean and CoV) of 7 VCI participants obtained during different walking conditions (unperturbed, sway1 and sway2; sway1: pseudorandom mediolateral platform sway with scaling factor A=0.005; sway 2: pseudorandom mediolateral platform sway, scaling factor A=0.0075 [see Figure3]) under different stimulation modes (Reference, A, B and C; see Figure 2). Each data point is derived from the final 100 steps of each condition.

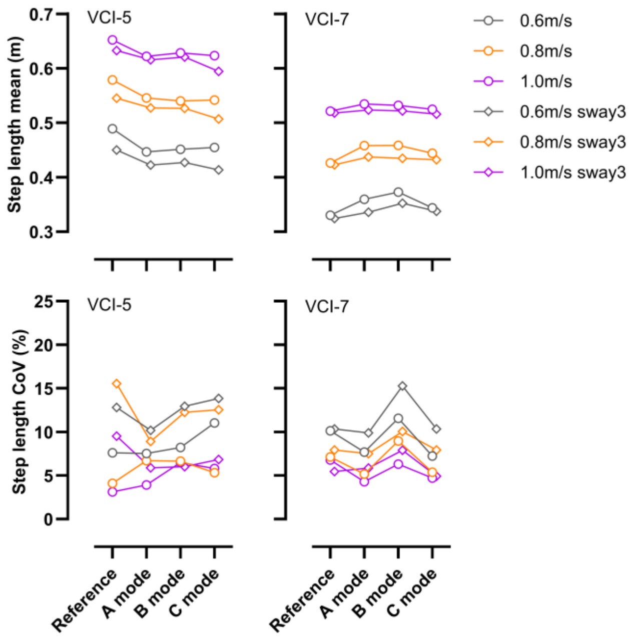


**eFigure 2**. Step length (mean and CoV) of 2 VCI participants obtained during different walking conditions (unperturbed, sway 3: pseudorandom mediolateral platform sway, scaling factor A=0.025 [see Figure3]) under different stimulation modes (Reference, A, B and C; see Figure 2). Each data point is derived from the final 100 steps of each condition.

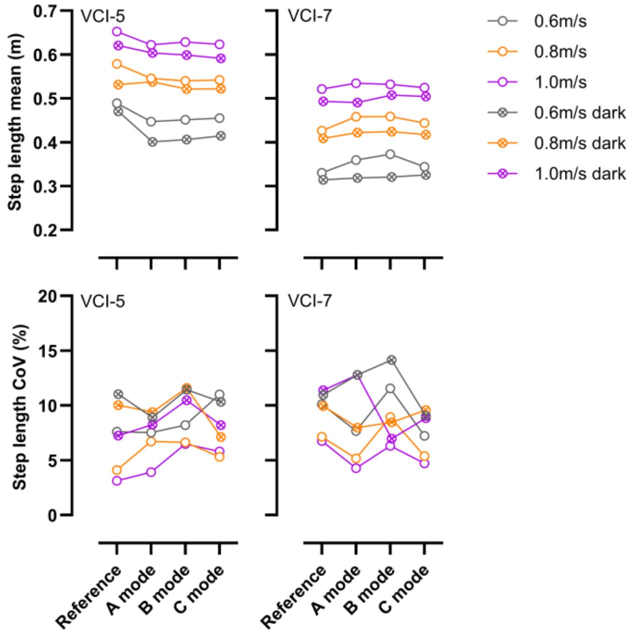


**eFigure 3**. Step length (mean and CoV) of 2 VCI participants obtained during different walking conditions (unperturbed, darkness) under different stimulation modes (Reference, A, B and C; see Figure 2). Each data point is derived from the final 100 steps of each condition.

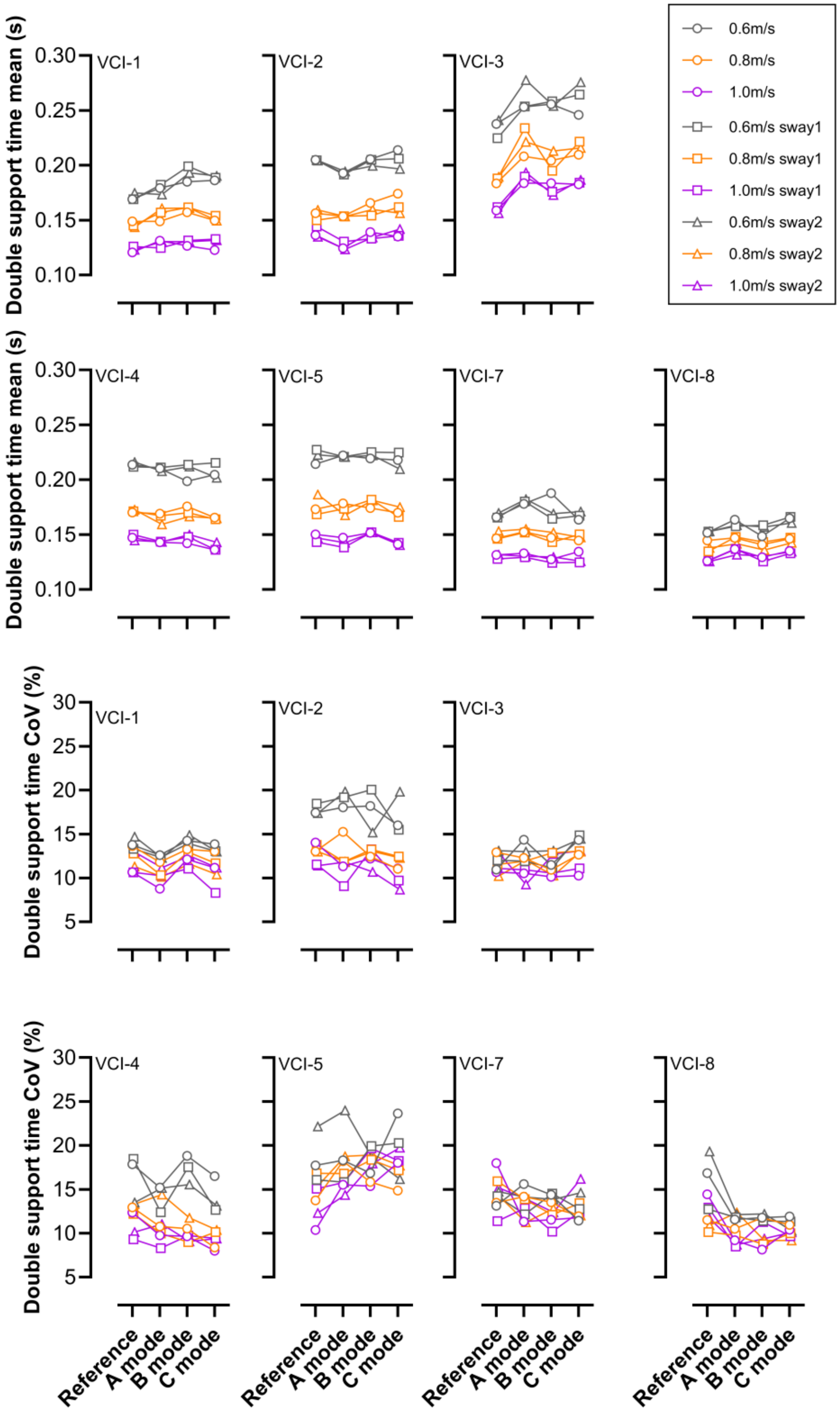


**eFigure 4**. Double support time (mean and CoV) of 7 VCI participants obtained during different walking conditions (unperturbed, sway1 and sway2; sway1: pseudorandom mediolateral platform sway with scaling factor A=0.005; sway 2: pseudorandom mediolateral platform sway, scaling factor A=0.0075 [see Figure3]) under different stimulation modes (Reference, A, B and C; see Figure 2). Each data point is derived from the final 100 steps of each condition.

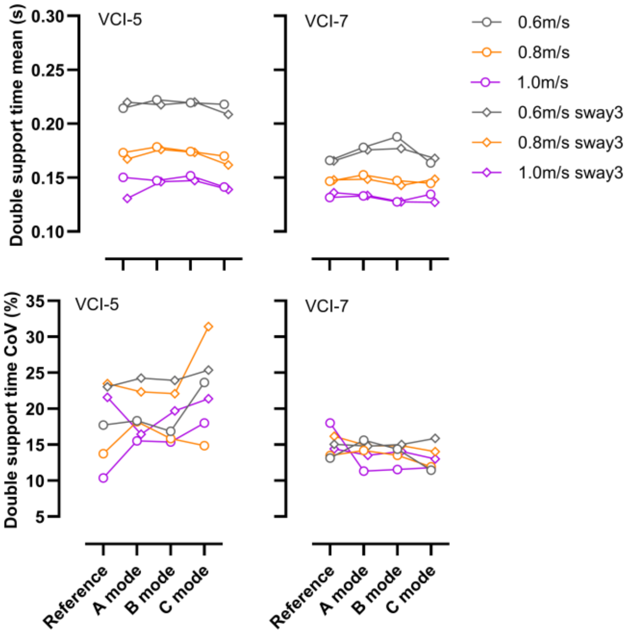


**eFigure 5**. Double support time (mean and CoV) of 2 VCI participants obtained during different walking conditions (unperturbed, sway 3: pseudorandom mediolateral platform sway, scaling factor A=0.025 [see Figure3]) under different stimulation modes (Reference, A, B and C; see Figure 2). Each data point is derived from the final 100 steps of each condition.

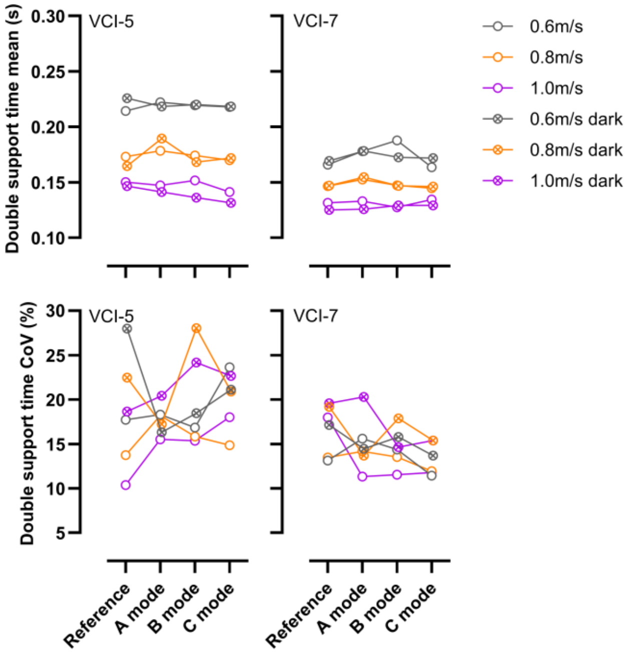


**eFigure 6**. Double support time (mean and CoV) of 2 VCI participants obtained during different walking conditions (unperturbed, darkness) under different stimulation modes (Reference, A, B and C; see Figure 2). Each data point is derived from the final 100 steps of each condition.

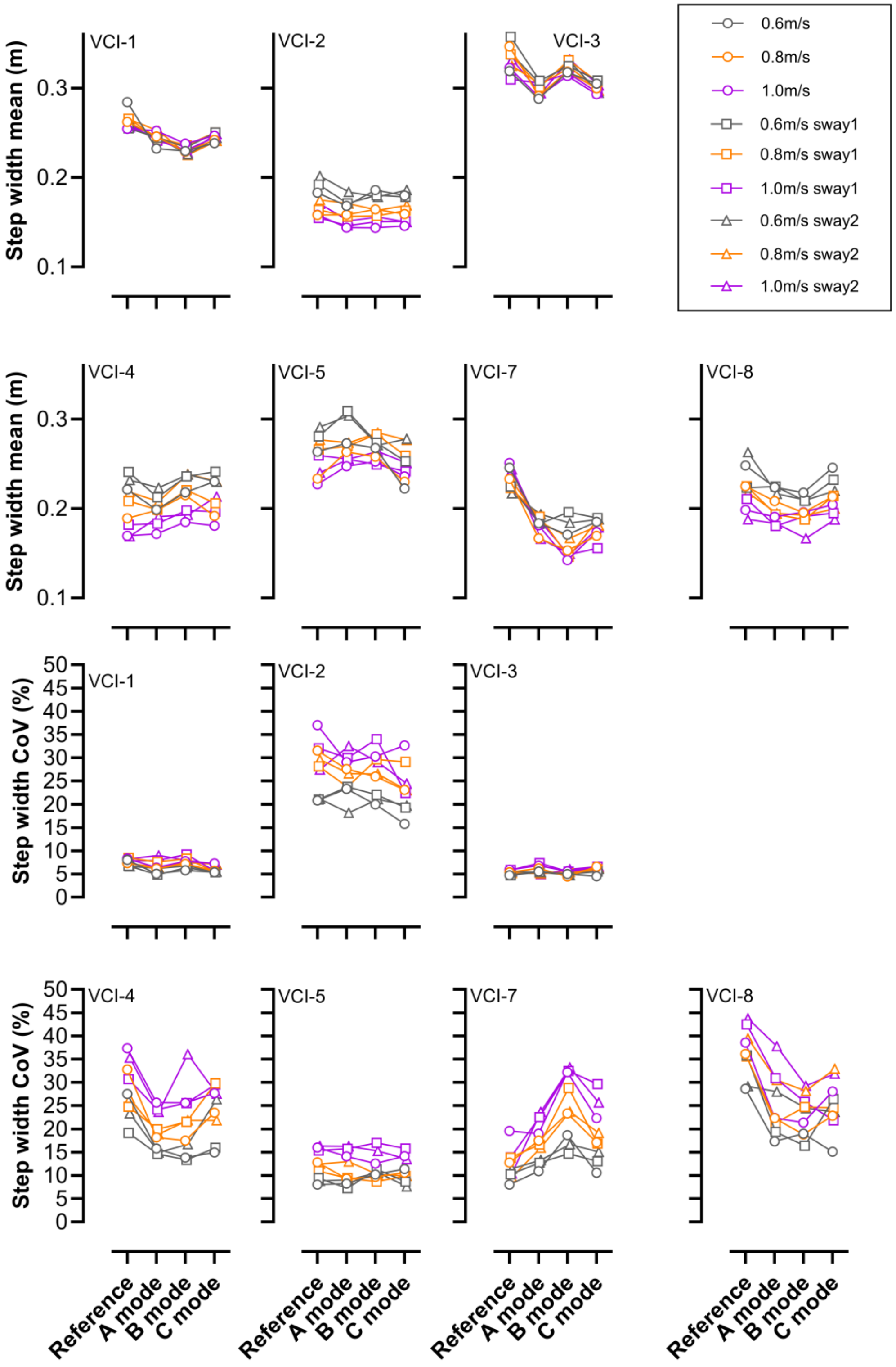


**eFigure 7**. Step width (mean and CoV) of 7 VCI participants obtained during different walking conditions (unperturbed, sway1 and sway2; sway1: pseudorandom mediolateral platform sway with scaling factor A=0.005; sway 2: pseudorandom mediolateral platform sway, scaling factor A=0.0075 [see Figure3]) under different stimulation modes (Reference, A, B and C; see Figure 2). Each data point is derived from the final 100 steps of each condition.

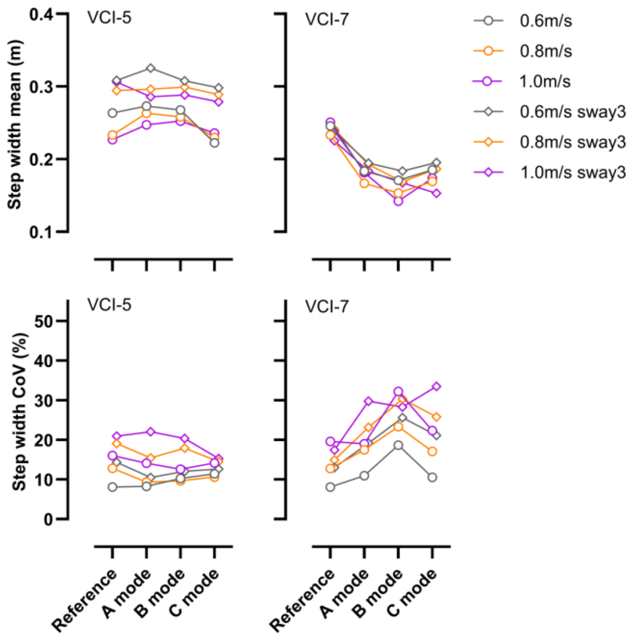


**eFigure 8**. Step width (mean and CoV) of 2 VCI participants obtained during different walking conditions (unperturbed, sway 3: pseudorandom mediolateral platform sway, scaling factor A=0.025 [see Figure3]) under different stimulation modes (Reference, A, B and C; see Figure 2). Each data point is derived from the final 100 steps of each condition.

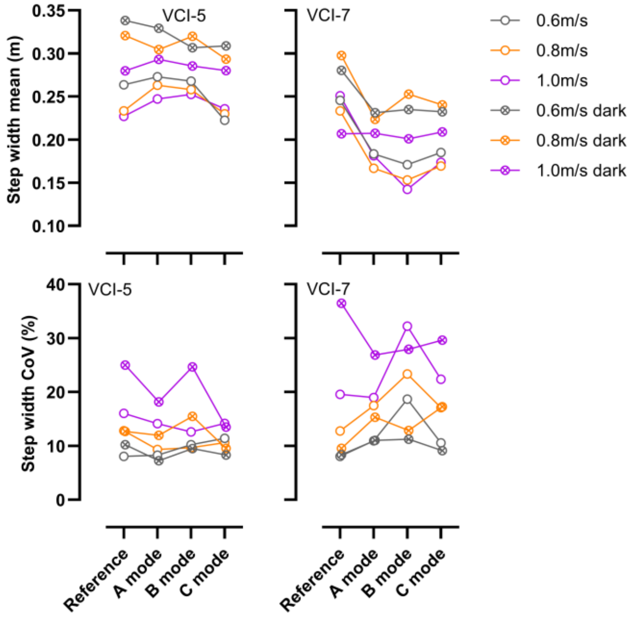


**eFigure 9**. Step width (mean and CoV) of 2 VCI participants obtained during different walking conditions (unperturbed, darkness) under different stimulation modes (Reference, A, B and C; see Figure 2). Each data point is derived from the final 100 steps of each condition.

*Exploratory Analyses: Patients with and without DFNA9*

The DFNA9 mutation, an autosomal dominant hereditary condition caused by mutations in the COCH gene, is known to be associated with vestibular dysfunction[1,2]. The presence of this mutation was not part of our original research questions in this study but 4 out of the 9 patients were known to carry this mutation and these participants have shown reduced responses to vestibular implant stimulation in A, B and C stimulation modes (lower eye velocities [see Figure 3, e.g., VCI-1 and VCI-4, in van Boxel, et al. [3] and movement perception [unpublished observations] when stimulating the implant). Given its potential impact, we explored the potential influence of the DFNA9 mutation on the primary outcome step time CoV, as well as the Mini-BESTest scores in the form of change scores, with positive values meaning a beneficial change in the stimulation modes compared to the reference value (lower variability and higher Mini-BESTest scores, respectively). Change in step time CoV was calculated such that positive values reflected reduced variability under stimulation (indicating improvement). As described above, it could be expected that the change scores would be smaller in DFNA9 carriers, reflecting less responsiveness to stimulation. As can be seen in eFigure 10, no consistent pattern across all walking speeds and conditions is apparent. During unperturbed walking, the median change scores in DFNA9 carriers appear to be similar or more positive compared to non-carriers and this pattern also appears in sway1 at 1.0 m/s and sway2 at 0.8 and 1.0 m/s. However, the opposite pattern is seen in sway1 at 0.6 and 0.8 m/s and sway2 at 0.6 m/s. In six of the nine walking conditions, the C mode appears to lead to the biggest difference, but the direction of the difference varies. Variation between participants is typically high (eFigure 10). Regarding the change in the Mini-BESTest scores, there were no clear differences between the change scores (eFigure 11), which also reflects the lack of a consistent pattern seen in Figure 7 (main manuscript) in the individual data.

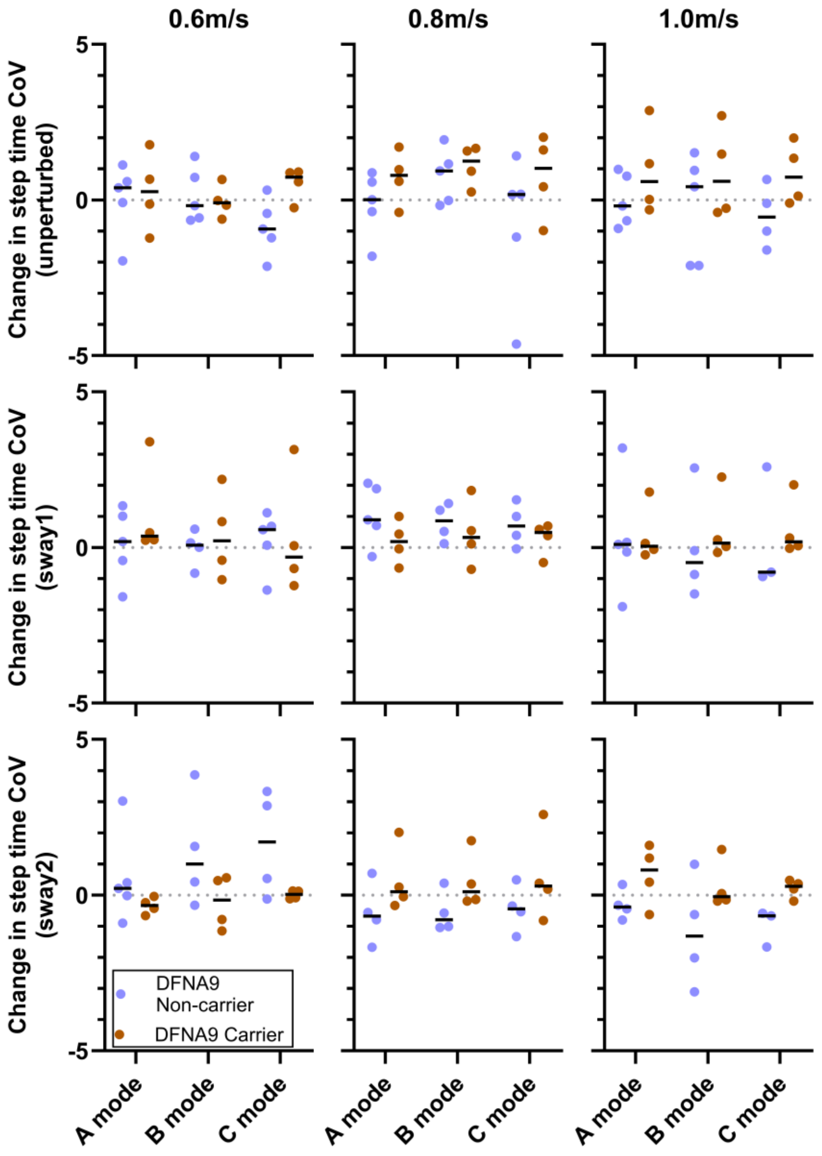


**eFigure 10**. Change in the step time CoV (individual values and group medians) from the reference condition to the stimulation conditions (A, B and C) during the unperturbed, sway1 and sway2 walking conditions at three walking speeds (0.6, 0.8 and 1.0 m/s) in participants carrying and not carrying DFNA9. Change was calculated such that positive change values indicate improvement (reduced variability).

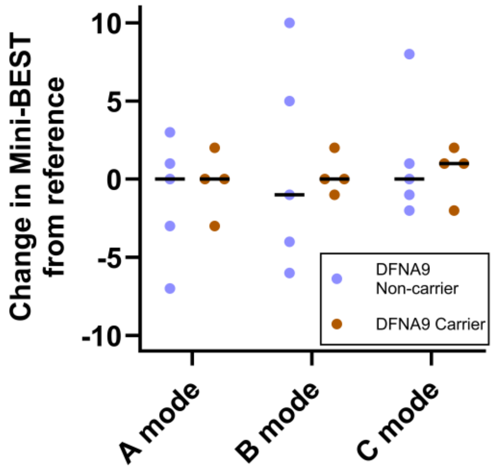


**eFigure 11**. Change in the Mini-BESTest total score (individual values and group medians) from the reference condition to the best of the two tests during stimulation periods (A, B and C) in participants carrying and not carrying DFNA9. Change was calculated such that positive change values indicate improvement (higher Mini-BESTest total score).

This lack of a clear pattern may be attributed to the small sample size or the high between-individual variability, which limits the ability to detect clear differences between the two groups. Alternatively it may be due to DFNA9 not particularly negatively impacting gait and balance performance and therefore not showing a clear difference in response under stimulation. To explore this possibility further, we visually compared the step time CoV results from the same unperturbed, sway1 and sway2 walking conditions from our previous study on people with BVP[4], separating the group into diagnosed DFNA9 carriers (n=7) and those with no known diagnosis (n=35) (eFigure 12). As shown in the figure, the DFNA9 carriers' values overlap with the no diagnosis group. While the small sample of seven carriers limits interpretation, the medians of the DFNA9 carriers tend to be below those of the non-carriers, which is contrary to the expectation that they would perform worse. This tentatively aligns with our suggestion above that the effects of DFNA9 might not particularly negatively impact gait and balance performance.

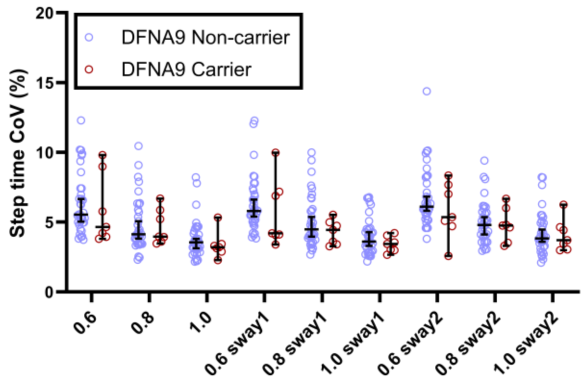


**eFigure 12**: Step time CoV (individual data, group medians and 95% confidence intervals) obtained during different walking conditions (no sway, sway1 and sway2) at three walking speeds (0.6, 0.8 and 1.0 m/s) from people with BVP derived from the data published in Zhu, et al. [4] separated into diagnosed DFNA9 carriers (n=7) and those with no known DFNA9 diagnosis (n=35). Each data point is derived from the final 100 steps of each condition.
